# The effect of transcranial direct current stimulation (tDCS) combined with cognitive training on EEG spectral power in adolescent boys with ADHD: a double-blind, randomised, sham-controlled trial

**DOI:** 10.1101/2021.07.21.21260953

**Authors:** Samuel J. Westwood, Natali Bozhilova, Marion Criaud, Sheut-Ling Lam, Steve Lukito, Sophie Wallace-Hanlon, Olivia S. Kowalczyk, Afroditi Kostara, Joseph Mathew, Bruce E. Wexler, Roi Cohen Kadosh, Philip Asherson, Katya Rubia

**Author notes:** Shared first-authorship and corresponding authors: Dr Samuel Westwood, Department of Child and Adolescent Psychiatry - PO85, Institute of Psychiatry, Psychology and Neuroscience, King’s College London, 16 De Crespigny Park, London, SE5 8AF, Dr Natali Bozhilova, School of Psychology, Elizabeth Fry Building, University of Surrey, Guildford, GU2 7XH UK.

## Abstract

Transcranial direct current stimulation (tDCS) is a possible neurotherapeutic alternative to psychostimulants in Attention-Deficit/Hyperactivity Disorder (ADHD). However, very little is known regarding the mechanisms of action of tDCS in children and adolescents with ADHD. We conducted the first multi-session, sham-controlled study of anodal tDCS over right inferior frontal cortex (rIFC), a consistently under-functioning region in ADHD, combined with cognitive training (CT) in 50 children/adolescents with ADHD. This study investigated the underlying mechanisms of action on resting and Go/No-Go Task-based QEEG measures in a subgroup of 23 participants with ADHD (n, sham=10; anodal tDCS=13). We found no significant sham versus anodal tDCS group differences in QEEG spectral power during rest and Go/No-Go Task performance, no correlation between the QEEG and Go/No-Go Task performance, and no effect on clinical and cognitive outcome measures. These findings extend the null clinical or cognitive effects in our whole sample of 50 children/adolescents with ADHD. Our findings do not indicate multi-session anodal tDCS with CT over rIFC as a treatment for children/adolescents with ADHD. Larger RCTs should explore different protocols titrated to the individual and using comprehensive measures to assess cognitive, clinical, and neural effects of tDCS and its underlying mechanisms of action in ADHD.

## Introduction

Attention-Deficit/Hyperactivity Disorder (ADHD) is neurodevelopmental disorder marked by age-inappropriate, and impairing symptoms of inattention and/or impulsivity-hyperactivity[1]. ADHD is also associated with deficits in executive functions (EF), including motor and interference inhibition, sustained attention, switching, working memory (WM), and timing[2], underpinned by neurofunctional abnormalities in inferior and dorsolateral fronto-striatal and fronto-cerebellar regions based on functional magnetic resonance imaging (fMRI) meta-analyses[3–6]. This atypical frontal brain activity in ADHD is further related to an increase in slow-wave cortical activity, as reflected in excessively increased electroencephalographic (EEG) power in theta and delta over frontal and central brain regions in both adults and children with ADHD[7,8].

The gold-standard treatment for ADHD are psychostimulants, which improve ADHD symptoms in roughly 70% of individuals with ADHD[9]. However, psychostimulants have been associated with side-effects[10], poor adherence in adolescence[11,12], while evidence of longer-term efficacy is limited[12,13], possibly due to brain adaptation[14]. Meta-analyses of alternative treatments, such as behavioural therapies, cognitive training (CT), or dietary interventions[15], result in small to moderate improvement in ADHD symptoms.

A promising neurotherapeutic alternative is transcranial direct current stimulation (tDCS), which can potentially modulate key dysfunctional brain regions associated with ADHD with longer-term neuroplastic effects that drugs cannot offer[2,16–18]. TDCS involves applying a weak direct electrical current via two electrodes (one anode, one cathode) placed on the scalp, which modulate the excitability of underlying brain regions via polarity-dependent, subthreshold shifts in resting membrane potentials. The net increase or decrease in neuronal excitability (under the anode or cathode, respectively) can modulate neuronal network activity[19], with these effects persisting after stimulation due to practice-dependent changes in synaptic plasticity, mediated by GABA, glutamate [20,21], dopamine, and noradrenaline[22–24]. Furthermore, unlike other forms of non-invasive brain stimulation, such as transcranial magnetic stimulation, tDCS is cheaper, easier to use, and well tolerated with minimal side effects[25].

Systematic reviews and meta-analyses of tDCS studies in ADHD suggest limited evidence of clinical or cognitive improvement with tDCS[26–28]. However, the majority of studies applied 1 to 5 tDCS sessions over mainly left dorsolateral prefrontal cortex (DLPFC), and only one study applied 5 sessions over the right inferior frontal cortex (IFC) [29]. No sham-controlled study so far has stimulated the IFC, a region most consistently shown to be under-functioning in ADHD[5,6], over 15 sessions combined with cognitive training (CT) with the aim of potentiating cognitive and clinical effects[30–35]. Thus, in the largest randomized, sham-controlled tDCS trial (RCT) in children and adolescents with ADHD, were applied sham or anodal tDCS over rIFC combined with cognitive training in EF across 15 consecutive weekday sessions in 50 children and adolescents with ADHD[36]. While both groups improved, we found no group differences in improvements in clinical symptoms or in cognitive performance (including motor and interference inhibition, sustained attention and vigilance, time estimation, visuo-spatial WM, and cognitive flexibility) immediately after treatment or at a 6-month follow-up [36].

Hardly anything is known about the neurophysiological substrates of tDCS effects in ADHD, with only three studies investigating these. In a double-blind, crossover RCT study with 10 adolescents with ADHD, single-session conventional anodal tDCS, anodal High Definition (HD)-tDCS or sham tDCS over rIFC led to enhanced N2 and P3 amplitude during an n-back WM task compared to sham[37]. In a double-blind RCT, 37 adults with ADHD received single sessions of sham or anodal tDCS over left or right DLPFC in a crossover design, with 18 participants performing an Eriksen Flanker task and 19 performing a Stop Signal task[38]. Participants showed reduced reaction times following left DLPFC and increased P3 amplitude following right and left DLPFC compared to sham in the Eriksen flanker task only, suggesting evidence of improved interference, but not response inhibition[38]. Using a functional cortical network (FCN) analysis on EEG activity, 50 adults with ADHD in a sham-control RCT showed increased functional brain connectivity within the stimulated and correlated areas after single-session anodal tDCS over the left DLPFC compared to baseline but not sham[39], thus we cannot rule out whether this improvement from baseline was incidental or a result of anodal tDCS specifically[39]. Given the scarcity of neurophysiological investigations in ADHD following tDCS, the present study investigated the mechanism of action of tDCS using EEG spectral power during rest and during a Go/No-Go motor inhibition task.

Compared to event-related potentials (ERPs), EEG spectral power has been a preferred measure of treatment/stimulation response in both clinical and non-clinical studies. Findings in healthy adults on the effects of tDCS on spectral power are mixed. In one RCT, single-session anodal tDCS over the rIFC led to a reduction in absolute theta power at rest and improved inhibitory performance compared to sham[40], suggesting that theta power might be the neural signature of successful post-treatment inhibition[40]. Further, compared to sham, RCTs with single-session anodal tDCS has also been shown to reduce frontro-central theta when stimulating left DlPFC[41] or enhanced theta-gamma coupling when stimulating right PFC[42]. EEG mean frequency was also found to be significantly reduced after both anodal and sham tDCS over the left DLPFC, although the effects were smaller for sham tDCS[43]. By contrast, more recent RCTs found no effects on both rest- and task-based EEG power spectrum following anodal tDCS[44–46], supporting a quantitative review that indicated little-to-no reliable neural effects of tDCS beyond motor evoked potentials (MEP)[47], although these findings might be due to small sample sizes and diverse methodology (e.g., differential measures and protocols) leading to discrepancy across non-clinical studies[48].

To our knowledge, this is the first study to examine the neuromodulatory effects of multi-session anodal tDCS combined with cognitive training over the rIFC on EEG spectral power in children and adolescents with ADHD. Based on aforementioned findings in healthy adults, we hypothesised that 15 sessions of anodal versus sham tDCS over the rIFC combined with multi-EF training would lead to a decrease during rest and an increase during task performance in absolute theta power. We also hypothesized that this effect would be associated with improved performance during a motor response inhibition Go/No-Go task.

## MATERIALS & METHOD

### Design

In a double-blind, sham-controlled, parallel RCT (ISRCTN: 48265228), 50 boys with ADHD received 15 sessions of anodal or sham tDCS over the rIFC combined with multi-EF training over 3 weeks[36]. We measured ADHD symptoms and related behaviours, ADHD-relevant EF (including motor and interference inhibition, sustained attention & vigilance, time estimation, working memory, and cognitive flexibility), safety, and EEG outcome measures at baseline, post-treatment, and 6-month follow-up. A more detailed experimental design can be found elsewhere[36]. Briefly, across 15 consecutive weekdays, participants received 20-minutes of 1mA anodal or sham tDCS over the rIFC (F8; cathode over right supra-orbital area, Fp1) while playing cognitive training games composed of ACTIVATE^TM^ games (to train visuo-spatial WM, selective attention, switching, and inhibition) and a training version of the Stop Task (to train motor inhibition)[36] Sham tDCS was identical to anodal tDCS except the current was administered for 60s (i.e., a 30s fade-in/fade-out)[36].

### Participants

For the purposes of this paper, only participants that completed baseline and post-treatment EEG recordings were included. This is because, of the 50 participants, 21 had no EEG data recorded at all, while only 16 participants had EEG data recorded at the 6-month follow-up, which was too few for data analysis. Thus, this left 13 participants in the anodal and 16 in the sham tDCS group with baseline and post-treatment EEG eligible for data analysis.

Twenty-nine male participants (10- to 18-years) had a clinical DSM-5 diagnosis of ADHD assessed by an experienced child psychiatrist and confirmed using the Schedule of Affective Disorders and Schizophrenia for School Age Children Present and Lifetime version (K□SADS□PL)[36]. Participants also had to score above cut-off on Conners 3^rd^ Edition–Parent Rating Scale (Conners 3-P, cut-off t-score > 60)[36], and were screened for Autism Spectrum Disorders (ASD) using the parent-rated Social Communication Questionnaire (SCQ, cut-off > 17)[36] and the pro-social scale of the Strengths & Difficulties Questionnaire (SDQ, cut-off < 5)[36]. Participants were excluded with IQsD□<80 (Wechsler abbreviated scale of intelligence, WASI-I)[36], a history of alcohol or substance abuse, neurological illness, comorbid major psychiatric disorders (except Conduct Disorder [CD]/Oppositional Defiant Disorder [ODD]); and tDCS contraindications. Consent was obtained from either the legal caregiver for participants under 16-years or from participants over 16-years (Figure 1). Participants received £540 for participating and were reimbursed for travel expenses.

Baseline assessment was scheduled at least two weeks after medication titration. Eighteen participants received stable ADHD medications (non-psychostimulants: 4; psychostimulants: 14; between 3-weeks and 9-years). To minimise the risk that psychostimulants might mask the effect of stimulation, participants on psychostimulants were asked to abstain for at least 24 hours before each assessment session. Of the 12 participants who abstained, 5 chose to abstain throughout the trial period (>24 hours before baseline until after post-assessment), while 7 abstained 24 hours before each the baseline and post-assessment only (see Table 1).

**Table 1:** Baseline demographic, medication, clinical, and cognitive measures; the number of tDCS and CT sessions; and the time spent playing each CT game in the sham and anodal tDCS groups

|  | Anodal tDCS<br>N=10 |  | Sham tDCS<br>N=13 |  | Independent t-test |  |
| --- | --- | --- | --- | --- | --- | --- |
| Demographics | Mean | SD | Mean | SD | t(2, 21) | p |
| *Age (months)* | 154.30 | 23.03 | 174.07 | 22.38 | -2.07 | 0.05 |
| *IQ (WASI-II)* | 104.50 | 15.40 | 106.77 | 13.73 | -0.37 | 0.71 |
| *Years in education* | 8.40 | 2.01 | 10.15 | 1.82 | -2.19 | 0.04 |
| *SCQ* | 9.60 | 5.582 | 8.62 | 6.826 | 0.37 | 0.72 |
| *SDQ (Prosocial)* | 7.00 | 2.055 | 6.31 | 1.974 | 0.82 | 0.42 |
| *Kiddie-SADS (ADHD Symptoms)* |  |  |  |  |  |  |
| Combined | 12.40 | 3.44 | 12.15 | 2.70 | 0.19 | 0.85 |
| Inattention | 7.60 | 1.27 | 7.69 | 0.95 | -0.20 | 0.84 |
| Hyperactivity/impulsivity | 4.80 | 2.70 | 4.46 | 2.47 | 0.31 | 0.76 |
| **Clinical Measures & Side Effects** |  |  |  |  |  |  |
| *ADHD-Rating Scale* |  |  |  |  |  |  |
| Total Score | 44.10 | 7.520 | 38.31 | 6.033 | 2.05 | 0.05 |
| Inattention Subscale | 24.60 | 2.413 | 21.46 | 3.799 | 2.28 | 0.03 |
| Imp/Hyp Subscale | 19.50 | 6.133 | 16.85 | 3.738 | 1.29 | 0.21 |
| *Conners 3-P (T-Score)* |  |  |  |  |  |  |
| ADHD Index | 17.20 | 2.044 | 14.92 | 3.662 | 1.76 | 0.09 |
| Global Index | 86.30 | 4.692 | 82.92 | 8.549 | 1.12 | 0.27 |
| DSM-5 Inattention | 87.60 | 2.716 | 82.15 | 7.221 | 2.26 | 0.04 |
| DSM-5 Hyp/Imp | 85.10 | 6.350 | 83.15 | 10.367 | 0.52 | 0.61 |
| *ARI* |  |  |  |  |  |  |
| Parent-rated | 0.97 | 0.55 | 0.83 | 0.53 | 0.60 | 0.56 |
| Child-rated | 0.70 | 0.53 | 0.66 | 0.39 | 0.24 | 0.82 |
| *MEWS* | 17.50 | 6.10 | 16.54 | 7.86 | 0.32 | 0.75 |
| *WREMB-R Total Score* | 23.10 | 6.95 | 20.54 | 4.03 | 1.11 | 0.28 |
| *CIS* | 23.40 | 9.69 | 22.00 | 6.60 | 0.41 | 0.68 |
| *Side effects* | 16.80 | 12.90 | 12.15 | 5.97 | 1.15 | 0.26 |
| **Cognitive Measures** |  |  |  |  |  |  |
| *GNG (PI %)* | 37.25 | 17.01 | 53.46 | 19.41 | -2.093 | .049 |
| *CPT* |  |  |  |  |  |  |
| Omissions % | 18.33 | 9.13 | 12.05 | 14.15 | 1.219 | .236 |
| Commissions % | 2.98 | 2.02 | 1.941 | 3.94 | .755 | .459 |
| *Simon Task (Simon RT Effect)* | 99.87 | 34.304 | 67.92 | 31.37 | 2.326 | .030 |
| *Time Discrimination Task (Total Correct)* | 66.17 | 10.45 | 79.74 | 14.37 | -2.514 | .020 |
| *Macworth Clock Task* |  |  |  |  |  |  |
| Omissions % | 49.25 | 13.13 | 30.00 | 16.99 | 2.962 | .01 |
| Commissions % | 10.56 | 12.24 | 2.20 | 1.72 | 2.447 | .023 |
| *WCST* |  |  |  |  |  |  |
| Perseverative Errors | 13.50 | 4.33 | 14.00 | 5.12 | -.248 | .807 |
| Non-Perservative Errors | 9.30 | 4.45 | 7.00 | 3.96 | 1.310 | .204 |
| *NIH Working Memory Task* | 19.60 | 16.37 | 31.92 | 9.74 | -2.254 | .035 |
| *Verbal Fluency (% Correct)* | 89.75 | 10.47 | 93.28 | 7.89 | -.924 | .366 |
| *Semantic Fluency (% Correct)* | 93.58 | 5.25 | 95.50 | 5.57 | -.842 | .409 |
| **Cognitive Training** |  |  |  |  |  |  |
| Grub Ahoy | 21.00 | 8.10 | 26.92 | 12.17 | -1.33 | 0.20 |
| Magic Lens | 58.50 | 14.54 | 58.85 | 10.64 | -0.07 | 0.95 |
| Monkey Trouble | 23.00 | 10.60 | 22.31 | 14.52 | 0.13 | 0.90 |
| Peter's Printer Panic | 102.00 | 12.52 | 90.39 | 17.73 | 1.76 | 0.09 |
| Treasure Trunk | 62.00 | 19.18 | 66.15 | 17.34 | -0.54 | 0.95 |

| **Medication** | *N* | *N* | $\chi^2$ (4) | *p* |
| --- | --- | --- | --- | --- |
| Medication-naïve | 1 | 3 | 2.54 | .64 |
| On-medication | 1 | 0 |  |  |
| On-medication, abstained for assessments | 2 | 5 |  |  |
| Off-medication | 3 | 2 |  |  |

ADHD-RS, Caregiver-rated ADHD Rating Scale;
ARI, Affective Reactivity Index;
CIS, Columbia Impairment Scale Parent;
Kiddie Schedule for affective disorders and schizophrenia present and lifetime version DSM-5 for ADHD;
MEWS, Mind Excessively Wandering Scale;
SD, Standard Deviation;
SCQ, Social Communication Questionnaire;
SDQ, Social Difficulties Questionnaire;
WASI-II, Wechsler Abbreviated Scale of Intelligence;

### Outcome Measures

#### Offline cognitive measures

The adult version of the Maudsley Attention and Response Suppression (MARS) Task battery[36] was used to measure motor response inhibition (Go/No-Go Task; dependent variable [DV]: % probability of inhibition [PI]), sustained attention (Continuous Performance Task [CPT]; DV: omission and commission errors), interference inhibition (Simon Task; DV: Simon reaction time effect), and time discrimination (Time Discrimination Task; DV: percentage correct). Other tasks measured vigilance (The Mackworth Clock Task[49–51]; DV: percentage omissions and commission errors), cognitive flexibility (Wisconsin Card Sorting Task, WCST[52]; DV: total and perserverative errors), visuo-spatial WM (C8 Sciences version of the NIH List Sorting Working Memory Task[53]; DV: total score). Given the possibility of a downregulation of left IFC mediated functions, particularly language production, verbal and semantic fluency (DV: percentage correct responses)[54] were also measured (a full description of tasks, is provided in Westwood et al[36]).

#### ADHD Symptoms and related impairments

Treatment effects in ADHD symptoms was measured with the caregiver-rated ADHD Rating Scale–IV (ADHD-RS) Home Version (DV: Total Scores)[55] and Conners 3-P (DV: ADHD Index)[56]. Also measured were related difficulties and functional impairments (Weekly Parent Ratings of Evening and Morning Behaviour-Revised scale, WREMB-R[57]; Columbia Impairment Scale-Parent version, CIS[58]); irritability (child- and caregiver-rated Affective Reactivity Index, ARI[59]), and mind-wandering (child-rated Mind Excessively Wandering Scale, MEWS)[60].

#### Safety measures

Safety was measured with caregiver-rated side effects[61] and adverse events[62].

### EEG-task description

Participants performed one block of the adult variant of the Maudsley Attention and Response Suppression (MARS) Go/No-Go Task, a measure of motor response inhibition[63,64]. In 73.4% of trials, a spaceship (Go stimulus) pointing left appeared in the centre of the screen and participants had to press the left with their left-index finger arrow key as fast as possible. In 26.6% of trials, a blue planet (No-Go stimulus) appeared in the centre of the screen instead of a spaceship and participants had to inhibit their response. Go and No-Go stimuli were displayed for 300ms followed by a blank screen for 1000ms. There are 150 trials in total (110 Go trials, 40 No-Go trials). The key dependent measure of the inhibitory performance is the probability of inhibition (PI). For completeness we also report other measures such as premature responses to all trials which is another impulsiveness indicator and the executive go process which includes mean reaction times (MRT), intrasubject response variability (i.e., SD of MRT) and omission errors to go trials. The task duration was 2.5 min. EEG was recorded over 9 minutes, with a 1-minute gap between rest and task activity, 5-mins resting activity, and ∼2.5-mins for task-related activity in the same order for all participants.

### EEG system/device

EEG was recorded from an 8-channel DC-coupled recording system using a wearable headset, manufactured by gtec (using Nautilus platform, https://www.gtec.at). Active dry electrodes (Sahara) with gold-plated pins and pre-amplification module were attached to the cap system, which allowed recordings using the 10-20 montage. The EEG data was wirelessly (using Bluetooth) transmitted to the recording laptop.

### EEG recording

During all recordings, the pre-amplification module in the active electrodes allows to keep the signal stable (∼20µV) and the impedances below (30kΩ). Adhesive ground and Ω reference electrodes were positioned at the mastoids. The signal was digitised at a sampling rate of 500 Hz, with additional online filters (bandpass filter, 0.1-100Hz, and notch filter, 58Hz-62Hz).

Participants were seated on a height adjustable chair in a testing lab. Stimuli were presented on a laptop at a distance of approximately 30cm.

### EEG pre-processing

Analyses were carried out in the open-source EEGLAB software[65]. Researchers were blind to group status during EEG pre-processing, analysis and discussion. The raw EEG data were re-referenced offline to the average reference and were down-sampled to 256 Hz. The raw data were also digitally filtered using basic Finite Impulse Response (FIR) filters between 0.1Hz and 30 Hz. Prior to re-referencing, flat channels and channels with extremely large artifacts were removed and replaced with topographic spline interpolation. On average, 2 channels were interpolated across all datasets. Sections of data exceeding 200μV were automatically removed. Ocular artifacts were removed using the independent component analysis (ICA) algorithm, runica, [66]. All other components were back-projected for further analysis. Following the back-projection, all datasets were also visually inspected and sections of data containing residual artefacts were removed manually. All analyses included EEG recordings which had 25% or less data removed, with 150-210 epochs of artifact free data on average (∼60 epochs for task performance, ∼150 epochs for rest episodes).

### QEEG

Quantitative EEG was investigated for resting state and EEG-Go/No-Go Task. Data were segmented into 2s epochs and power spectra were computed using a fast Fourier transform with a 10% Hanning window. Analyses focused on alpha (8–14 Hz), theta (3–7 Hz) and beta (15–30 Hz) band differences between two groups (anodal vs sham tDCS). EEG absolute power density (μV²/Hz) within each frequency bands was averaged across all electrode sites (Cz, F7, F3, F4, F8, Fpz, Fz, Pz) and the entire recording duration to reduce the number of statistical comparisons. The same analysis was performed for F8 alone; as this electrode was the stimulation site. To further explore treatment-related change, theta activity was also calculated by subtracting theta activity at post-treatment from EEG activity during baseline (Supplementary Analysis 3).

### Statistical analysis

Normality of data was assessed using the Shapiro–Wilk statistic and visual inspection of score distribution. Log 10 transformation of the EEG data and error data was performed to normalise both the EEG and the error data. Furthermore, exploratory pairwise correlational analysis among age, ADHD severity, all EEG and cognitive performance measures were performed and reported in the Supplementary Material. The correlational results indicated that younger age at entry was moderately associated with higher theta and alpha activity during rest at baseline and during task performance at post-treatment, as well as with greater intra-subject variability, slower reaction time and more premature errors during the QEEG Go/No-Go at both baseline and post-treatment (Supplementary Table 2). Greater ADHD severity was also moderately associated with poorer Go/No-Go PI. There were no other significant correlations (Supplementary Table 2).

Group differences on all outcome measures were tested with repeated measures analysis of covariance (ANCOVA) with Group (anodal vs sham tDCS) as a between-subjects factor and Time (baseline vs post-treatment) as a within-subjects factor, while covarying for baseline age in months and ADHD-RS scores. The covariates were selected to adjust for baseline differences, with the anodal tDCS group being significantly younger and reported higher ADHD-RS Total Score (see Table 1).

The alpha level was set at 0.05. To correct for multiple testing, False Discovery Rate (FDR) correction with Benjamini-Hochberg[67] was applied to outcomes with *p*-values less than 0.1, which was applied separately to the different frequency bands, secondary clinical outcomes, and secondary cognitive outcomes. We did not correct for multiple testing on Offline Go/No-Go, or ADHD-RS as these were considered primary outcome measures (see also Westwood et al., 2021[36]). In the Results section below, we report significant *p*-values before and after FDR correction (hereafter referred to as unadjusted and FDR adjusted, respectively). Analyses were conducted with IBM SPSS Statistics 26 (IBM Corp., Armonk, N.Y., USA).

### Ethics

This trial received local research ethics committee approval (REC ID: 17/LO/0983) and was conducted in accordance with the Declaration of Helsinki [68].

## RESULTS

Out of the 29 individuals, 6 individuals were excluded from all analyses due to extreme values driven by large EEG artifacts. Only 23 recordings were available for the analysis (n=10 active, n=13 sham).

### Baseline Comparisons

Compared to sham, the anodal tDCS group was on average 2-years younger, had higher ADHD-RS Total & Inattentive Scores and Conners’ 3-P DSM-Inattentive Scores, and fewer years in education. Cognitively, the anodal compared to the sham tDCS group showed significantly poorer performance on the following offline tasks: Go/No-Go (PI%), Simon (Simon RT Effect), Macworth Clock (Omissions & Commissions), and NIH WM Tasks (Total Score) (see Table 1).

### EEG Outcomes Measures

#### During Rest

There was no significant Group-by-Time interaction and no significant main effects of Group or Time on EEG activity (alpha, beta, theta) during rest based on the average of all electrodes or at F8 only (Table 2).

**Table 2.**
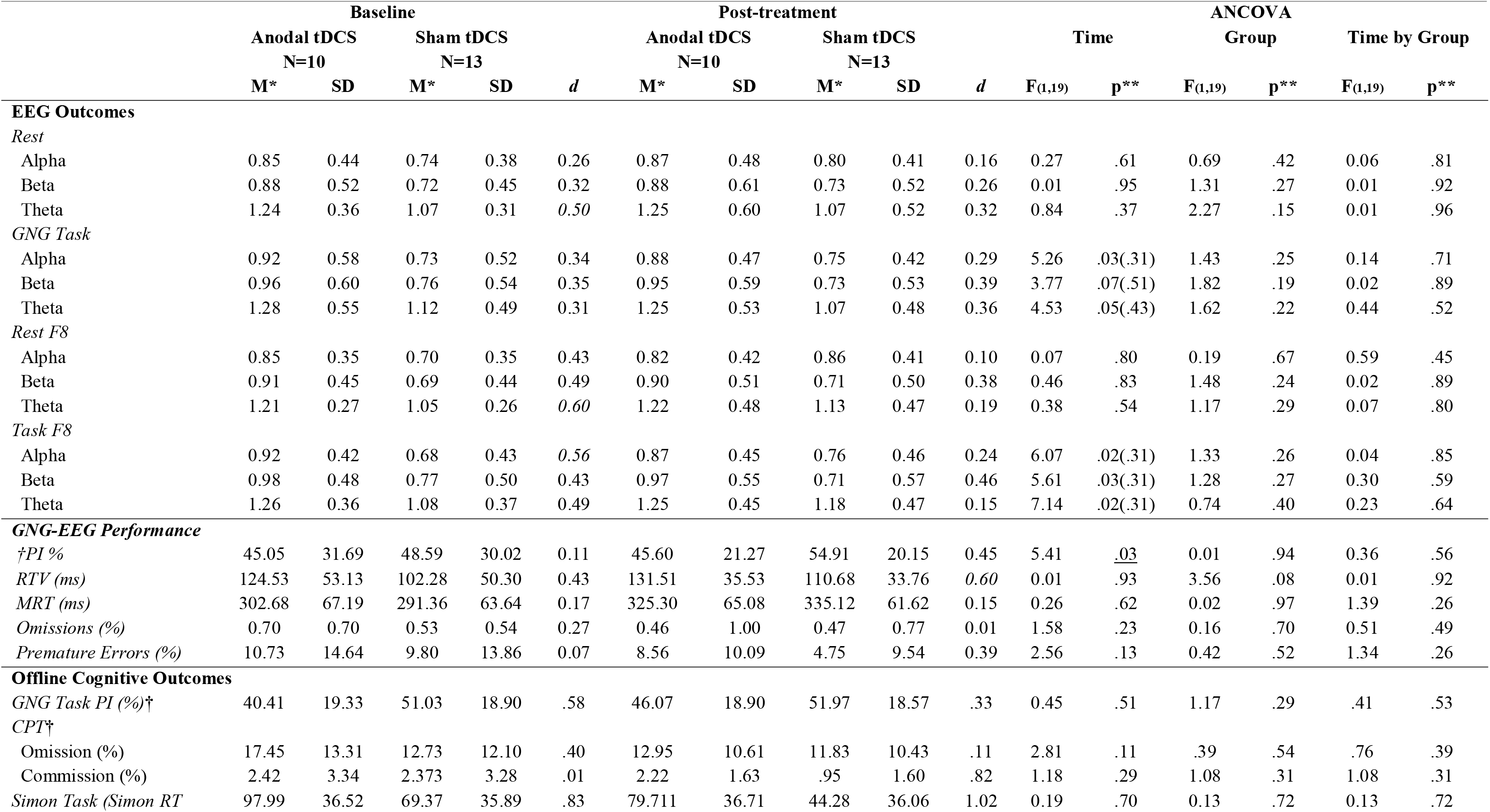
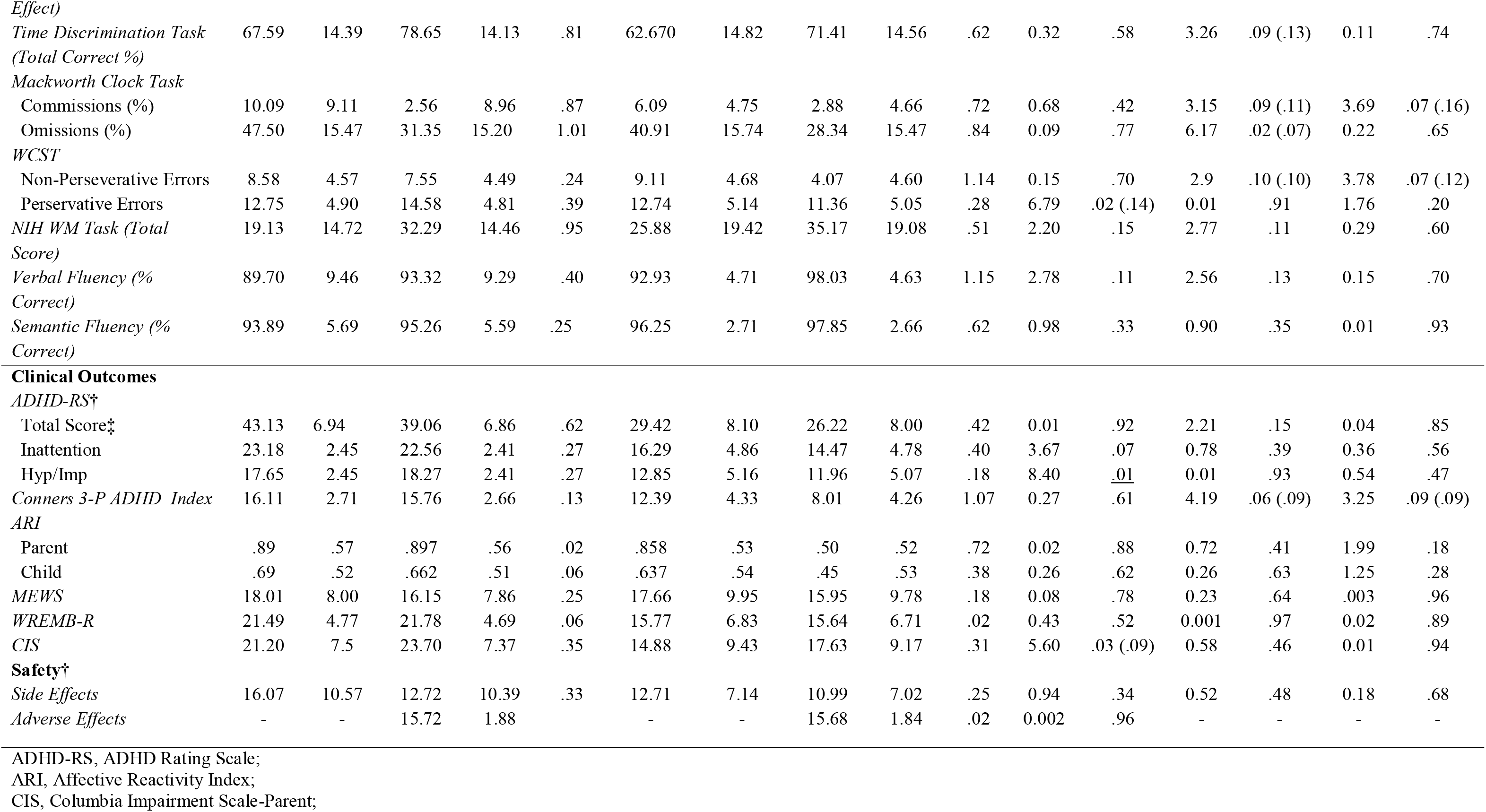
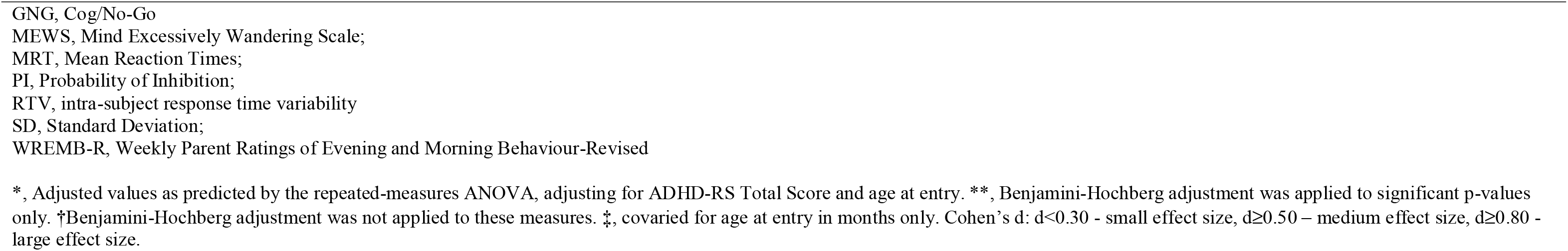
Summary of adjusted average performance on EEG outcomes, primary and secondary cognitive and clinical outcome measures after sham and anodal tDCS combined with CT. Benjamini-Hochberg adjusted p-values given parentheses.

#### During Go/No-Go Task Performance

Based on the average across all electrodes, there was no significant Group-by-Time interaction effect and no significant main effect of Group on EEG activity during Go/No-Go Task performance. There was a main effect of Time on theta and alpha activity, with lower theta and alpha activity at post-treatment compared to baseline, but this effect did not survive FDR correction (Table 2). There was a significant time-by-age interaction (theta, F(1,19)=6.72, unadjusted p=0.018, FDR adjusted p= 0.31; alpha, F(1,19)=5.42,unadjusted p=0.032, FDR adjusted p= 0.31) showing higher theta and alpha activity in younger subjects at pre- compared to post-treatment, but this interaction was no longer significant after FDR correction.

Based on F8 only, there was no significant Group-by-Time interaction effect and no significant main effect of group on EEG activity (alpha, beta, theta). There was a significant main effect of Time, showing lower alpha, beta and theta activity at post-treatment compared to baseline, but this was no longer significant after FDR correction. There was also a significant time-by-age interaction effect (theta, F(1,19)=9.91,unadjusted p=0.006, FDR adjusted p=0.10; alpha, F(1,19)=6.39,unadjusted p=0.021, FDR adjusted p=0.31; beta (F(1,19)= 5.86, unadjusted p=0.026, FDR adjusted p=0.31), showing higher theta and alpha activity in younger subjects at pre- compared to post-treatment, which was no longer significant after FDR correction (Table 2).

#### Other Clinical & Offline Cognitive Outcome Measure

There was no significant main effects of Group, Time, or Group-by-Time interaction effect that survived FDR Correction on clinical or offline cognitive measures (Table 2), except a significant main effect of Time in ADHD-RS Hyperactivity/Impulsivity Subscale (F(1,19)=8.4, *p*=0.01).

## DISCUSSION

This is the first double-blind, sham-controlled RCT to test the neurofunctional mechanisms of action of multi-session, anodal tDCS over the rIFC combined with cognitive training in children and adolescents with ADHD on QEEG spectral power. Sham and anodal tDCS did not differ on QEEG spectral power during rest and Go/No-Go Task performance. Further, both sham and real tDCS showed lower EEG spectral power at post-treatment compared to baseline, although this did not survive FDR correction for multiple comparisons. Finally, there were no differences in these subgroups in clinical and cognitive outcome measures. The absence of group differences in QEEG spectral power, and the pattern of results in clinical and cognitive outcomes in this subgroup of 23 children and adolescents with ADHD extends the evidence of overall null clinical or cognitive effects of anodal tDCS over the rIFC relative to sham in the original sample of 50 children and adolescents with ADHD published elsewhere[36]. The lack of QEEG effects may relate to the lack of clinical and cognitive benefits of tDCS by showing no tDCS-related electrophysiological effects as measured in EEG measures during rest and a cognitive control task. These null findings are also complemented by the lack of correlation between the QEEG and Go/No-Go cognitive performance measures in the same subgroups. However, the null EEG findings need to be seen with caution considering the small sample.

The lack of any tDCS effect on QEEG measures contrasts with previous evidence of reduced absolute theta power during rest after a single session of anodal compared to sham tDCS over the rIFC in adults without ADHD[40]. Another study in 15 adolescents with ADHD found enhanced ERP amplitudes (e.g., N2 and P3)[37,38] during an n-back WM task after a single-session of conventional anodal tDCS or HD-tDCS over rIFC. Nevertheless, our findings support studies that used more fine-grained measures of EEG activity and failed to identify differences between anodal and sham tDCS over the left DLPFC in EEG connectivity during rest in 50 adults with ADHD[39], or any effect on both rest- and task-based EEG power spectrum following anodal tDCS in neurotypical adults[44–46]. Together, our findings extend a quantitative review showing little-to-no reliable neurophysiological effects of anodal tDCS in neurotypical adults[69].

These findings might suggest that multi-session tDCS over rIFC combined with CT had no effect on EEG measures of spectral power during rest or task performance in children with ADHD. However, this lack of effect on EEG spectral power could be related to the low sample size (n=23) and/or baseline differences, with the anodal tDCS group being younger and having higher parent ratings on ADHD-RS and Conners’ 3-P compared to sham. A potential confounding effect of age chimes with previous evidence indicating that QEEG varies as a function of age[70]. Children with ADHD consistently show excessively high absolute delta and theta power compared to adults with ADHD, and this excessive increase in slow frequencies decreases with age and hence becomes less different from neurotypical individuals [7,8]. Furthermore, with the original sample of 50 children with ADHD, older but not younger participants showed less improvement in ADHD symptoms in the anodal versus sham tDCS group at post-treatment[36]. This finding might be explained by less ADHD severity in the older compared to the younger participants at baseline. However, differences in clinical measures and age were covaried for. Future studies should investigate how age affects EEG activity following anodal tDCS in individuals with and without ADHD.

Both groups showed improvement in the EEG Go/No-Go PI and offline WCST Perseverative Errors from baseline to post-treatment. A similar effect of time effect was observed in the larger sample of 50 ADHD children for cognitive performance on an offline Go/No-Go and other offline tasks measuring ADHD-related EF (e.g., Simon RT Effect; Verbal Fluency)[36]. However, in the absence of a Time-by-Group interaction, the baseline to post-treatment improvement in QEEG Go/No-Go PI and WCST Perseverative Errors could either be due to CT, to placebo and/or to practice effects. The correlational analysis showed a negative correlation between EEG Go/No-Go PI with ADHD severity (ADHD-RS total), but not with CT and EEG measures, suggesting the improvement in EEG Go/No-Go PI was not related to CT or EEG changes. However, we cannot rule out this is a chance result given the small sample of participants analysed. Together, findings from the EEG Go/No-Go Task and offline cognitive tasks broadly replicate cognitive findings in the whole group[36].

Given the insufficient EEG data at the 6-month follow-up assessment, we could not test for potential longer-term tDCS effects on EEG measures after a period of consolidation[71]. However, thus far, tDCS studies in ADHD that show longer-term clinical and/or cognitive effects have all reported significant effects at post-treatment that persisted only in the order of weeks, not months[72,73]. This study found no post-treatment effects, and the analysis of the whole group found no clinical or cognitive effects at follow-up[36], making longer-term effects in EEG measures unlikely. Future studies should investigate consolidation effects in EEG measures following anodal tDCS in children with ADHD, which – at the time of writing – has not been studied. Other neurotherapies, such as EEG neurofeedback (NF) or fMRI-NF[74], have shown stronger clinical effects at follow-up than at post-treatment, indicating a delayed consolidation of neuromodulatory effects, suggesting neuroplasticity [74–76]. Unfortunately, we only collected EEG data of 16 participants at follow-up, and it was hence not possible to investigate the longer-term effects of anodal tDCS on EEG activity in the current study. Future studies should therefore focus on evaluating the effect of tDCS on neural activity at multiple follow-up points (e.g., 3 and 9 months) and using better spatially resolved techniques such as fMRI.

### Limitations

Although this RCT had a larger sample (n=50) than other tDCS studies in children and adolescents with ADHD, EEG data could only be collected for 29 participants with only 23 analysed, and there was insufficient EEG data at follow-up (n=16) to test longer-term effects of tDCS. The findings (based on 23 participants) are hence underpowered and will need to be replicated using larger sample sizes. The attrition rate of this study was very high (42%) compared to studies using QEEG in children above the age of 4 either with and without ADHD (5%-25%)[77–79], which is likely due to the discomfort of the dry EEG electrodes[80] that were chosen for their ease of application. Future studies should aim at choosing electrodes, which induce the minimum possible discomfort in children and adolescents with ADHD. Technological advances mean that dry active EEG electrode can reliably estimate EEG spectral power and ERP components, but they still suffer high interelectrode impedance and therefore very high noise levels[81], as evidenced in our data and in the exclusion of 6 participants as outliers. Future studies with dry electrodes should consider including tasks with longer duration (15 minutes) and a sufficient number of trials (100 or more trials), which should boost statistical power and provide more confidence estimates of tDCS effects. Additionally, this study did not include EEG triggers to capture the stimuli presentation and had only eight electrodes, resulting in a limited choice of analyses. Therefore, we cannot exclude the possibility that event-related analyses might have been more informative and led to positive findings. Future studies should take advantage of both the time and frequency domain of EEG, and thus perform various event-related time-frequency analyses. The use of 64 or more electrodes would also allow for EEG source-level analysis, which could be invaluable in understanding the exact cortical origin of the EEG signal.

### Conclusions

This study in 23 children with ADHD did not show a differential effect of 15 sessions of anodal versus sham tDCS & CT on EEG spectral power. Furthermore, there were also no cognitive or clinical benefits of tDCS & CT in this subgroup. The findings extend our previous findings in a larger group of 50 ADHD children of no superior effects of anodal versus sham tDCS & CT on clinical or cognitive measures by showing no underlying neurofunctional mechanism of action in a subgroup[36]. Although tDCS is becoming increasingly accepted into clinical practice and viewed as an alternative to medication by parents [17,82,83], our findings suggest that rIFC stimulation may not be indicated as a treatment choice for neurophysiological, cognitive or clinical remediation for children and adolescents with ADHD. Larger RCTs need to be conducted to explore different protocols (such as different stimulation sites, amplitude, frequency, etc) titrated to the individual and using cognitive, clinical, and neural outcome measures to comprehensively assess the effect of tDCS and its underlying mechanisms of action on brain activity in ADHD.

## Supporting information

Supplemental Analysis

## Data Availability

Data is available upon reasonable request

## Supplementary Materials

The supplementary material for this article can be found in Supplementary Analysis.docx

## Acknowledgements

We would like to thank the families and children who took part in this study, and the South London and Maudsley NHS Trust and local parent groups for their support and participation in this study. We would like to thank Yuanyuan Yang for her help in recruiting participants and data collection. Finally, we thank Professor Bruce Wexler and Sean O’Leary for granting us access to and for providing free IT support with the ACTIVATE™.

## Author Contributions

Contributions according to Contributor Roles Taxonomy (CRediT) (casrai.org/credit/)

|  |  |
| --- | --- |
| Conceptualisation | Samuel J. Westwood, Natali Bozhilova, Marion Criaud, Sheut-Ling Lam, Steve Lukito, Bruce Wexler, Roi Cohen-Kadosh, Philip Asherson and Katya Rubia |
| Data curation | Samuel J. Westwood, Natali Bozhilova, Marion Criaud, Sheut-Ling Lam, Steve Lukito, Sophie Wallace-Hanlon and Afroditi Kostara |
| Formal analysis | Samuel J. Westwood, Natali Bozhilova, Roi Cohen-Kadosh, and Katya Rubia |
| Funding acquisition | Bruce Wexler, Roi Cohen-Kadosh, Philip Asherson and Katya Rubia |
| Investigation | Samuel J. Westwood, Marion Criaud, Sheut-Ling Lam, Steve Lukito, Sophie Wallace-Hanlon, Olivia S. Kowalczyk, Afroditi Kostara and Joseph Mathew |
| Methodology | Samuel J. Westwood, Marion Criaud, Sheut-Ling Lam, Steve Lukito, Olivia S. Kowalczyk, Bruce Wexler, Roi Cohen-Kadosh, Philip Asherson and Katya Rubia |
| Project administration | Samuel J. Westwood and Katya Rubia |
| Resources | Bruce Wexler, Roi Cohen-Kadosh, Philip Asherson and Katya Rubia |
| Software | Marion Criaud, Bruce Wexler and Katya Rubia |
| Supervision | Samuel J. Westwood and Katya Rubia |
| Validation | Samuel J. Westwood and Katya Rubia |
| Visualisation | Natali Bozhilova |
| Writing – original draft preparation | Samuel J. Westwood, Natali Bozhilova, and Katya Rubia |
| Writing – review and editing | Samuel J. Westwood, Natali Bozhilova, Marion Criaud, Sheut-Ling Lam, Steve Lukito, Sophie Wallace-Hanlon, Olivia S. Kowalczyk, Bruce Wexler, Roi Cohen-Kadosh and Katya Rubia |

## Institutional Review Board Statement

The authors assert that all procedures contributing to this work comply with the ethical standards of the relevant national and institutional committees on human experimentation and with the Helsinki Declaration of 1975, as revised in 2008. This trial received research ethics committee approval from The NHS Research Authority, London Camberwell St Giles Research Ethics Committee (REC ID: 17/LO/0983) in November 2017.

## Informed Consent Statement

Informed consent was obtained from all participants involved in the study.

## Data Availability Statement

Data is available upon reasonable request

