## Supplemental Analysis for "The effect of transcranial direct current stimulation (tDCS) combined with cognitive training on EEG spectral power in adolescent boys with ADHD: a double-blind, randomised, sham-controlled trial"

**Supplementary Materials**

**Supplementary EEG Analysis 1: Individual Electrodes**

***Electrode, F3***

**EEG activity during Rest.** There were no significant group-by-time interaction, group or time effects on EEG activity (alpha, beta, theta) during rest (see Supplementary Table 1).

**EEG activity during Go/No-Go Task.** There was only a significant time -by-age interaction on theta activity (F(1,19)=6.19,p=0.020). Younger participants had higher EEG activity at baseline compared to older participants, suggesting the main effect of time might be primarily driven by age (see Supplementary Table 1).

***Electrode, F4***

**EEG activity during Rest.** There were no significant group-by-time interaction, group or time effects on EEG activity (alpha, beta, theta) during rest (see Supplementary Table 1).

**EEG activity during Go/No-Go Task.** There were no significant main effects of group or group-by-time interaction (Supplementary Table 1). There was a main effect of time on theta and alpha activity before adjusting for multiple comparisons. Theta and alpha activity were reduced at post-treatment compared to pre-treatment in both groups. There was also a significant time-by-age interaction (theta, F(1,19)=13.75,p=0.002; alpha, F(1,19)=11.95,p=0.003) after adjusting for multiple comparisons. Younger participants had higher EEG activity at pre-treatment compared to older participants, suggesting the main effect of time might be primarily driven by age.

***Electrode, F7***

**EEG activity during Rest.** There were no significant group-by-time interaction, group or time effects on EEG activity (alpha, beta, theta) during rest (see Supplementary Table 1).

**EEG activity during Go/No-Go Task.** There were no significant group-by-time interaction, group or time effects on EEG activity (alpha, beta, theta) during task (see Supplementary Table 1).

***Electrode, Fz***

**EEG activity during Rest**. There were no significant group-by-time interaction, group or time effects on EEG activity (alpha, beta, theta) during rest (see Supplementary Table 1).

**EEG activity during Go/No-Go Task.** There were no significant group-by-time interaction, group or time effects on EEG activity (alpha, beta, theta) during task performance (see Supplementary Table 1).

***Electrode, Cz***

**EEG activity during Rest**. There were no significant group-by-time interaction, group or time effects on EEG activity (alpha, beta, theta) during rest (see Supplementary Table 1).

**EEG activity during Go/No-Go Task.** There were no significant group-by-time interaction, group or time effects on EEG activity (alpha, beta, theta) during task performance (see Supplementary Table 1).

***Electrode, Pz***

**EEG activity during Rest**. There were no significant group-by-time interaction, group or time effects on EEG activity (alpha, beta, theta) during rest (see Supplementary Table 1).

**EEG activity during Go/No-Go Task.** There were no significant group-by-time interaction, group or time effects on EEG activity (alpha, beta, theta) during task performance (see Supplementary Table 1).

**Supplementary Analysis 2: Theta/Beta Ratio *with All Electrodes (F3, F4, F7, F8, Fpz, Fz, Pz, Cz)***

***Theta/Beta Ratio at Rest***

There were no significant group-by-time interaction, group or time effects on theta/beta ratio during rest (see Supplementary Table 1).

***Theta/Beta Ratio during Go/No-Go Task.***

There were no significant group-by-time interaction, group or time effects on theta/beta ratio during task performance (see Supplementary Table 1).

**Supplementary Analysis 3: Comparisons between responders and non-responders**

Responder status was defined as the presence of an increase in theta activity at post-treatment compared to pre-treatment. Non-responder status was defined as the absence of an increase in theta activity at post-treatment compared to pre-treatment.

 Repeated measures ANOVA revealed no main effect of response status (responder vs non-responder vs sham) on all clinical and EEG measures (F=3.20, p=0.061). Nevertheless, we conducted post-hoc analyses.

***Clinical measures***

Compared to responders, non-responders had higher baseline (ARI-child rated) scores (p=0.036) and lower scores on the Monkey Trouble cognitive training games scores (p=0.019).

***EEG measures***

Compared to responders, non-responders had higher theta (p=0.006 rest; p=0.001 task performance) and alpha (p=0.007 rest; p=0.002 task performance) activity during both rest and EEG Go/No-Go Task performance at baseline.

 	Responders showed greater treatment-related change in theta activity during rest (p=0.011) and task (p=0.038) compared to non-responders.

There were no significant differences between responders and non-responders on any task performance measures (EEG-Go/No-Go task or offline cognitive tasks).

**Supplementary Analysis 4: Correlations**

Pearson’s correlations were administered to investigate whether change scores (post-treatment minus baseline) in ADHD-RS or Go/No-Go were associated with changes in EEG measures. The results showed no significant correlation with changes in EEG measures with either ADHD-RS or Go/No-Go Task Performance (Supplementary Table 2).

| **Table 1.** Descriptive statistics, effect sizes and ANCOVA for each electrode | | | | | | | | | | | | | | | | |
| --- | --- | --- | --- | --- | --- | --- | --- | --- | --- | --- | --- | --- | --- | --- | --- | --- |
|  | **Baseline** | | | |  | **Post-treatment** | | | |  | **ANCOVA** | | | | | |
|  | **Active**  **N=10** | | **Sham**  **N=13** | |  | **Active**  **N=10** | | **Sham**  **N=13** | |  | **Time** | | **Group** | | **Time by Group** | |
|  | **M*** | **SD** | **M*** | **SD** | ***d*** | **M*** | **SD** | **M*** | **SD** | ***d*** | **F** | **p*** | **F** | **p*** | **F** | **p*** |
| ***Rest F3*** |  |  |  |  |  |  |  |  |  |  |  |  |  |  |  |  |
| Alpha | 0.88 | 0.31 | 0.60 | 0.30 | 0.91 | 0.86 | 0.41 | 0.81 | 0.40 | 0.12 | 0.01 | .95 | 2.47 | .13 | 0.90 | .35 |
| Beta | 0.88 | 0.34 | 0.60 | 0.33 | 0.84 | 0.86 | 0.48 | 0.81 | 0.47 | 0.11 | 0.03 | .86 | 1.61 | .22 | 0.40 | .54 |
| Theta | 1.32 | 0.28 | 0.98 | 0.27 | 1.24 | 1.21 | 0.45 | 1.10 | 0.44 | 0.25 | 0.01 | .94 | 3.69 | .07(.51) | 1.21 | .28 |
| ***Task F3*** |  |  |  |  |  |  |  |  |  |  |  |  |  |  |  |  |
| Alpha | 0.90 | 0.41 | 0.65 | 0.43 | 0.60 | 0.87 | 0.34 | 0.79 | 0.35 | 0.23 | 3.64 | .07(.51) | 1.71 | .21 | 0.62 | .44 |
| Beta | 0.91 | 0.41 | 0.72 | 0.42 | 0.46 | 0.90 | 0.42 | 0.78 | 0.43 | 0.28 | 1.38 | .26 | 1.11 | .31 | 0.10 | .76 |
| Theta | 1.29 | 0.43 | 0.93 | 0.44 | 0.83 | 1.19 | 0.35 | 1.12 | 0.36 | 0.20 | 3.82 | .07(.51) | 2.29 | .15 | 1.84 | .19 |
| ***Rest F4*** |  |  |  |  |  |  |  |  |  |  |  |  |  |  |  |  |
| Alpha | 0.83 | 0.28 | 0.66 | 0.28 | 0.61 | 0.78 | 0.43 | 0.78 | 0.42 | 0.01 | 0.39 | .54 | 0.53 | .48 | 0.63 | .44 |
| Beta | 0.91 | 0.32 | 0.63 | 0.32 | 0.88 | 0.84 | 0.50 | 0.73 | 0.49 | 0.22 | 0.62 | .44 | 1.72 | .21 | 0.54 | .47 |
| Theta | 1.21 | 0.22 | 1.00 | 0.22 | 0.95 | 1.17 | 0.47 | 1.04 | 0.47 | 0.28 | 0.12 | .74 | 1.97 | .18 | 0.13 | .72 |
| ***Task F4*** |  |  |  |  |  |  |  |  |  |  |  |  |  |  |  |  |
| Alpha | 1.31 | 0.28 | 0.99 | 0.29 | 1.12 | 1.14 | 0.41 | 1.07 | 0.43 | 0.17 | 9.43 | .03(.21) | 1.96 | .18 | 2.42 | .14 |
| Beta | 1.03 | 0.36 | 0.66 | 0.37 | 1.01 | 0.93 | 0.40 | 0.70 | 0.41 | 0.57 | 3.11 | .09(.63) | 4.09 | .06(.42) | 0.59 | .45 |
| Theta | 1.31 | 0.28 | 0.99 | 0.29 | 1.12 | 1.14 | 0.41 | 1.07 | 0.43 | 0.17 | 9.66 | .02(.21) | 2.14 | .16 | 2.06 | .17 |
| ***Rest F7*** |  |  |  |  |  |  |  |  |  |  |  |  |  |  |  |  |
| Alpha | 0.82 | 0.40 | 0.78 | 0.39 | 0.10 | 0.93 | 0.33 | 0.77 | 0.33 | 0.49 | 2.24 | .15 | 0.63 | .44 | 0.34 | .57 |
| Beta | 0.82 | 0.40 | 0.78 | 0.39 | 0.10 | 0.93 | 0.33 | 0.77 | 0.33 | 0.49 | 0.11 | .75 | 0.51 | .49 | 0.24 | .63 |
| Theta | 1.27 | 0.39 | 1.21 | 0.39 | 0.15 | 1.36 | 0.43 | 1.08 | 0.42 | 0.65 | 1.36 | .26 | 1.12 | .30 | 1.33 | .26 |
| ***Task F7*** |  |  |  |  |  |  |  |  |  |  |  |  |  |  |  |  |
| Alpha | 0.91 | 0.47 | 0.81 | 0.49 | 0.21 | 0.93 | 0.32 | 0.79 | 0.33 | 0.43 | 3.74 | .07(.51) | 0.63 | .44 | 0.04 | .84 |
| Beta | 0.94 | 0.53 | 0.79 | 0.54 | 0.28 | 0.96 | 0.39 | 0.78 | 0.40 | 0.45 | 1.23 | .28 | 0.86 | .37 | 0.01 | .93 |
| Theta | 1.28 | 0.44 | 1.17 | 0.45 | 0.25 | 1.37 | 0.37 | 1.15 | 0.39 | 0.58 | 2.24 | .15 | 1.15 | .30 | 0.34 | .57 |
| ***Rest Fz*** |  |  |  |  |  |  |  |  |  |  |  |  |  |  |  |  |
| Alpha | 0.76 | 0.45 | 0.75 | 0.44 | 0.02 | 0.85 | 0.35 | 0.75 | 0.35 | 0.29 | 0.14 | .71 | 0.11 | .74 | 0.28 | .61 |
| Beta | 0.81 | 0.51 | 0.73 | 0.53 | 0.15 | 0.92 | 0.40 | 0.66 | 0.41 | 0.64 | 0.04 | .84 | 0.89 | .36 | 0.74 | .40 |
| Theta | 1.09 | 0.41 | 1.07 | 0.40 | 0.05 | 1.22 | 0.43 | 1.05 | 0.42 | 0.40 | 1.00 | .33 | 0.38 | .55 | 0.79 | .39 |
| ***Task Fz*** |  |  |  |  |  |  |  |  |  |  |  |  |  |  |  |  |
| Alpha | 0.87 | 0.52 | 0.64 | 0.53 | 0.44 | 0.89 | 0.39 | 0.70 | 0.40 | 0.48 | 2.12 | .16 | 1.45 | .25 | 0.02 | .89 |
| Beta | 0.90 | 0.58 | 0.77 | 0.62 | 0.22 | 0.99 | 0.45 | 0.70 | 0.48 | 0.62 | 2.11 | .16 | 1.10 | .31 | 0.35 | .56 |
| Theta | 1.19 | 0.46 | 1.06 | 0.47 | 0.28 | 1.27 | 0.36 | 1.07 | 0.38 | 0.54 | 1.58 | .19 | 1.07 | .31 | 0.18 | .68 |
| ***Rest Cz*** |  |  |  |  |  |  |  |  |  |  |  |  |  |  |  |  |
| Alpha | 0.78 | 0.39 | 0.83 | 0.39 | 0.13 | 0.68 | 0.47 | 0.74 | 0.45 | 0.13 | 0.63 | .44 | 0.19 | .67 | 0.01 | .99 |
| Beta | 0.80 | 0.42 | 0.75 | 0.42 | 0.12 | 0.68 | 0.50 | 0.59 | 0.49 | 0.18 | 0.16 | .69 | 0.17 | .68 | 0.03 | .87 |
| Theta | 1.11 | 0.35 | 1.09 | 0.35 | 0.06 | 1.06 | 0.47 | 0.99 | 0.46 | 0.15 | 1.54 | .23 | 0.10 | .76 | 0.04 | .85 |
| ***Task Cz*** |  |  |  |  |  |  |  |  |  |  |  |  |  |  |  |  |
| Alpha | 0.85 | 0.46 | 0.80 | 0.47 | 0.11 | 0.64 | 0.42 | 0.57 | 0.43 | 0.16 | 1.48 | .24 | 0.13 | .73 | 0.01 | .94 |
| Beta | 0.88 | 0.43 | 0.76 | 0.44 | 0.28 | 0.67 | 0.42 | 0.52 | 0.41 | 0.36 | 3.66 | .07(.51) | 0.63 | .44 | 0.03 | .86 |
| Theta | 1.23 | 0.45 | 1.11 | 0.46 | 0.26 | 1.01 | 0.43 | 1.00 | 0.44 | 0.02 | 0.35 | .56 | 0.13 | .72 | 0.25 | .63 |
| ***Rest Pz*** |  |  |  |  |  |  |  |  |  |  |  |  |  |  |  |  |
| Alpha | 0.60 | 0.44 | 0.47 | 0.43 | 0.30 | 0.59 | 0.50 | 0.68 | 0.49 | 0.32 | 0.44 | .51 | 0.03 | .86 | 0.55 | .47 |
| Beta | 0.55 | 0.48 | 0.37 | 0.49 | 0.37 | 0.57 | 0.53 | 0.42 | 0.55 | 0.27 | 0.16 | .69 | 0.87 | .36 | 0.01 | .92 |
| Theta | 0.92 | 0.40 | 0.74 | 0.39 | 0.46 | 0.95 | 0.56 | 0.86 | 0.55 | 0.16 | 1.47 | .24 | 0.53 | .48 | 0.14 | .72 |
| ***Task Pz*** |  |  |  |  |  |  |  |  |  |  |  |  |  |  |  |  |
| Alpha | 0.64 | 0.51 | 0.45 | 0.53 | 0.37 | 0.56 | 0.37 | 0.50 | 0.38 | 0.16 | 3.14 | .09(.63) | 0.62 | .44 | 0.35 | .56 |
| Beta | 0.62 | 0.51 | 0.38 | 0.56 | 0.45 | 0.56 | 0.43 | 0.48 | 0.47 | 0.18 | 2.61 | .13 | 0.74 | .40 | 0.48 | .50 |
| Theta | 1.01 | 0.50 | 0.75 | 0.52 | 0.51 | 0.87 | 0.39 | 0.86 | 0.40 | 0.03 | 2.30 | .15 | 0.51 | .48 | 1.70 | .21 |

**Supplementary Table 2**. Correlations between age, ADHD-RS, EEG Frequency Bands, and Go/No-Go EEG measures

|  |  | **Age** | **ADHD-RS** | | | **Go/No-Go Measures** | | | | | **EEG measures** | | | | | |
| --- | --- | --- | --- | --- | --- | --- | --- | --- | --- | --- | --- | --- | --- | --- | --- | --- |
|  |  |  |  |  |  |  |  |  |  |  | Rest | | | Task Performance | | |
| **Age** | Years | * | *AI* | *HI* | *Total* | *MRT* | *RTV* | *PI* | *OE* | *Pre* | *Alpha* | *Beta* | *Theta* | *Alpha* | *Beta* | *Theta* |
| **ADHD-RS** | *AI* | -.11 | * |  |  |  |  |  |  |  |  |  |  |  |  |  |
|  | *HI* | -.20 | .74 | * |  |  |  |  |  |  |  |  |  |  |  |  |
|  | *Total* | -.16 | .93 | .93 | * |  |  |  |  |  |  |  |  |  |  |  |
| **Go/No-Go**  **measures** | *MRT* | -.20 | .06 | .13 | .10 | * |  |  |  |  |  |  |  |  |  |  |
|  | *RTV* | .05 | -.49 | -.44 | -.49 | -.02 | * |  |  |  |  |  |  |  |  |  |
|  | *PI* | -.15 | .45 | .56 | .54 | .52 | -.39 | * |  |  |  |  |  |  |  |  |
|  | *OE* | -.09 | .21 | -.01 | .10 | .16 | .20 | -.04 | * |  |  |  |  |  |  |  |
|  | *PrE* | .08 | .06 | .20 | .14 | -.24 | .13 | -.26 | -.34 | * |  |  |  |  |  |  |
| **EEG measures** | *Alpha* | .14 | .02 | -.25 | -.13 | -.16 | -.03 | -.08 | .11 | .29 | * |  |  |  |  |  |
|  | *Beta* | .03 | .10 | -.12 | -.02 | -.04 | -.14 | .02 | .11 | -.01 | .83 | * |  |  |  |  |
|  | *Theta* | .14 | .10 | -.16 | -.04 | -.20 | -.03 | -.15 | .12 | .32 | .92 | .69 | * |  |  |  |
|  | *Alpha* | -.22 | .05 | -.06 | -.01 | -.08 | -.10 | -.05 | .13 | -.14 | .72 | .91 | .54 | * |  |  |
|  | *Beta* | -.17 | .04 | -.01 | .02 | -.01 | -.10 | -.02 | .12 | -.22 | .64 | .90 | .44 | .97 | * |  |
|  | *Theta* | -.28 | -.01 | -.12 | -.07 | .01 | -.03 | -.08 | .18 | -.18 | .64 | .88 | .45 | .98 | .97 | * |

ADHD RS- ADHD Rating Scale IV, AI - Inattention, H- Hyperactivity, MRT – Mean Reaction Time, RTV- Reaction Time variability, PI- Probability of Inhibition, OE – Omission Errors PrE – Premature Errors, Red colour = significant at 0.05.
